# Preexisting diabetes and COVID-associated hyperglycaemia in patients with COVID-19 pneumonia

**DOI:** 10.1101/2021.04.17.21255548

**Authors:** Andrea Laurenzi, Amelia Caretto, Chiara Molinari, Elena Bazzigaluppi, Cristina Brigatti, Ilaria Marzinotto, Alessia Mercalli, Raffaella Melzi, Rita Nano, Cristina Tresoldi, Giovanni Landoni, Fabio Ciceri, Vito Lampasona, Marina Scavini, Lorenzo Piemonti

## Abstract

**Aim:** The aim of the current study was to compare clinical characteristics, laboratory findings and major outcomes of patients hospitalized for COVID-19 pneumonia with COVID-associated hyperglycaemia or preexisting diabetes.

**Methods:** A cohort of 176 adult patients with a diagnosis of pre-existing diabetes (n=112) or COVID-associated hyperglycaemia (n=55) was studied. Clinical outcomes and laboratory findings were analysed according to the presence of the two conditions. The time to viral clearance was assessed during the follow-up after hospital discharge.

**Result:** Patients with COVID-associated hyperglycaemia had lower BMI, significantly less comorbidities and higher levels of inflammatory markers and indicators of multi-organ injury than those with preexisting diabetes. No differences between preexisting diabetes and COVID-associated hyperglycaemia were evident for symptoms at admission, humoral response against SARS-CoV-2 or autoantibodies to glutamic acid decarboxylase or interferon alpha-4. COVID-associated hyperglycaemia was independently associated with the risk of adverse clinical outcome defined as ICU admission or death (HR 2.11, 95% CI 1.34-3.31; p=0.001), even after adjustment for age, sex and other selected variables associated with COVID-19 severity. Furthermore, we documented a negative association (HR 0.661, 95% CI 0.43-1.02; p=0.063) between COVID-associated hyperglycaemia and the time to swab negativization.

**Conclusions:** The recognition of hyperglycaemia as a specific clinical entity associated with COVID-19 pneumonia is relevant for early and appropriate patient management and close monitoring for the progression of disease severity.

## Introduction

Increasing evidence suggests the existence of a bidirectional link between COVID 19 and diabetes[1]. Preexisting diabetes is a risk factor for poor outcomes and death after SARS-CoV-2 infection[2-4]. On the other hand, new-onset hyperglycemia [5] and acute metabolic decompensation of preexisting diabetes [6] are now emerging as complications of COVID-19 pneumonia, especially in patients requiring hospitalization. This suggests a potential diabetogenic effect of SARS-CoV-2 infection, independent from the stress response to the acute illness[7]. Both reduced insulin secretion and increased insulin resistance have been proposed as possible mechanisms [8]. In fact, both endocrine and exocrine pancreatic tissues are susceptible to productive SARS-CoV-2 infection[9, 10], which may damage β-cell integrity, and decrease insulin secretion. Furthermore, the surge in cytokines production associated with the viral infection may be responsible for an increase in insulin resistance[11]. Regardless of the mechanism responsible for the metabolic dysregulation observed, the clinical entity of COVID-associated hyperglycaemia still has not been adequately characterized and separated from the entity of preexisting diabetes[5]. Few studies have comparatively characterized the two conditions as for the presence of comorbidities, pre-hospitalization treatments, symptoms at admission and laboratory variables associated with the severity of infection. Many studies have recently reported that COVID-associated hyperglycaemia is associated with a poorer outcome compared to that of the normoglycemic individuals[12-17]. However, whether COVID-associated hyperglycaemia is associated with a poorer clinical outcome compared to preexisting diabetes is still an open question, with conflicting findings [13, 15, 18-20]. Moreover, data on islet autoimmunity prevalence, anti SARS-CoV-2 antibody responses, and timing for viral clearance are still missing. Finally, it is unclear whether COVID-associated hyperglycaemia persists or reverts when the viral infection resolves. To address this gap in our knowledge, we studied a cohort of 176 adult patients with confirmed COVID-19 pneumonia, with a diagnosis of preexisting diabetes or hyperglycaemia.

## Material and Methods

### Study population and data sources

The study population consisted of 176 adult patients hospitalized for COVID-19 pneumonia with a diagnosis of preexisting diabetes or hyperglycaemia. These patients were selected from two previously described and partially overlapping cohorts [12], assembled at the IRCCS San Raffaele Hospital (Milan, Italy) as part of an institutional clinical– biological cohort (COVID-BioB; ClinicalTrials.gov Identifier: NCT04318366) of patients with COVID-19 pneumonia. The first cohort (n=584) was previously analysed for the humoral response against SARS-Cov-2 (n = 509)[21], while the second cohort (n=169) was studied for the development of thromboembolic complications. The study was approved by the local Institutional Review Board (Comitato Etico - Ospedale San Raffaele; protocol n 34/int/2020; NCT04318366) and all patients signed a written informed consent. A confirmed case was defined as presence of symptoms and radiological findings suggestive of COVID-19 pneumonia and/or a SARS-CoV-2-positive RT-PCR test from a nasal/throat (NT) swab. Demographic information, clinical features and laboratory tests were obtained within 72 hours from admission. Data were collected directly through patients’ interviews or medical chart review. Data regarding patients’ comorbidities were collected during hospitalization or after discharge from both structured baseline patient interviews and hospital paper or electronic medical records. All disease diagnoses present at the date of infection were included as comorbidities. Data were verified by data managers and clinicians for accuracy and crosschecked in blind. Data were recorded until hospital discharge or in-hospital death, whichever occurred first. We also recorded mortality and swab negativization beyond hospital discharge through our dedicate follow-up clinic: for patients non attending the follow-up clinic, we verified patient’s vital status with either family members or family physician.

### Laboratory variables

Routine blood tests encompassed serum biochemistry [including renal and liver function, lactate dehydrogenase (LDH)], complete blood count with differential, markers of myocardial damage [troponin T and pro-brain natriuretic peptide (proBNP)], and inflammation markers [C-reactive protein (CRP), ferritin, erythrocyte sedimentation rate (ESR), interleukin-6 (IL-6), procalcitonin]. An extended coagulation profile was obtained including D-dimer, PT, PTT, fibrinogen and antithrombin activity. Specific antibodies to different SARS-CoV-2 antigens, interferon alpha-4 and glutamic acid decarboxylase (GAD) were tested by a luciferase immunoprecipitation system (LIPS) assay, as previously described (11).

### Definition of diabetes

Study participants were defined as having: a) preexisting diabetes if they had a documented diagnosis of diabetes before the hospital admission for COVID-19 pneumonia [fasting plasma glucose (FPG) ≥7.0 mmol/l or HbA1c ≥ 6.5% (48 mmol/mol), or they were prescribed diabetes medications]; b) COVID-associated hyperglycaemia if they had a mean fasting plasma glucose in the absence of infusions of dextrose ≥7.0 mmol/l during the hospitalization for COVID-19 pneumonia in the presence of a negative history for diabetes and/or normal glycated hemoglobin level in the last year and in the absence of prescribed diabetes medications. We computed mean fasting glucose and glucose variability (standard deviation) from all fasting laboratory glucose values measured during hospitalization. The diagnosis of pancreatogenic and steroid-induced diabetes was attributed to patients whose diabetes was reasonably secondary to their exocrine pancreatic disease or steroid use. The diagnosis of type 1 diabetes was attributed to patients whose diabetes at onset was associated with evidence of islet autoimmunity. The diagnosis of type 2 diabetes was attributed to all patients who did not fulfil the criteria for any of the above.

### Statistical analysis

Categorical variables are reported as frequency or percent, continuous variables as median with inter-quartiles range (IQR) in parenthesis. Categorical variables were compared using Chi-square or Fischer’s exact test, as appropriate; continuous variables using the Mann-Whitney test. Associations between baseline variables and diabetes status was assessed by logistic regression. Multiple regression models, each adjusted for age and sex and one additional baseline variable of interest were performed. The effect estimates were reported as Odd Ratios (ORs), outcome variable were pre-existing diabetes ore new onset diabetes as appropriate to obtain OR >1.

Survival was estimated according to Kaplan–Meier. The time-to-event was calculated from the date of symptom onset to the date of the event, or of last follow-up visit, whichever occurred first. We used univariate and multivariate Cox proportional hazards models to study the association between patient characteristics with time to adverse outcome (a composite endpoint of admission to ICU or death, whichever occurred first). The effect estimates were reported as Hazard Ratios (HRs) with the corresponding 95% CI estimated according to the Wald approximation. Multivariate analyses were performed including variables significant at the level of <0.1 in the univariate analysis. Variables were excluded from multivariate Cox regression if they showed substantial biological redundancy with other variables (e.g., aspartate aminotransferase vs alanine aminotransferase), had data obtained for fewer than 78% of patients, in order to prevent model overfitting. Two-tailed P values are reported, with P value <0.05 indicating statistical significance. All confidence intervals are two-sided and not adjusted for multiple testing. Statistical analyses were performed with the SPSS 24 (SPSS Inc. /IBM) and the R software version 3.4.0 (R Core Team (2020).

## Results

### Study participants

The study population consisted of 176 adult patients (≥18 years) with pre-existing diabetes or COVID-associated hyperglycaemia admitted between February 25^th^ and May 2^nd^ 2020 to the Emergency or Clinical departments of the IRCCS San Raffaele Hospital (Milan, Italy). Among the 176 cases included in the study 11 (6.3%) were discharged without hospitalization, 137 (77.8%) were treated with non-invasive ventilation and 39 (22.2%) were admitted to the ICU over the hospitalization period. The median hospital stay was 17 (8-17) days. As of February 14, 2021 the median follow-up time after symptoms onset was 213 (95% CI: 199-227) days. Sixty-four patients died (36.4%). According to the composite endpoint of ICU admission or death, 82 patients (46.6%) had an adverse in-hospital outcome.

### COVID-associated hyperglycaemia was associated with specific basal characteristics

Preexisting diabetes and COVID-associated hyperglycaemia accounted for 68.8% (n=121) and 31.25% (n=55) of our cohort, respectively. Among the 121 subjects with pre-existing diabetes, 109 (90.1%) were diagnosed with type 2 diabetes, 4 (3.3%) with type 1 diabetes and 8 (6.6%) with secondary diabetes (5 pancreatogenic, 3 steroid-induced). The characteristics of the study participants for patients with pre-existing diabetes or COVID-associated hyperglycaemia at the time of hospitalization are reported in Table 1. Sex- and age-adjusted logistic regression analysis was used to assess the associations between baseline variables and pre-existing diabetes vs COVID-associated hyperglycaemia. Higher BMI at admission [after log1p transformation OR 19.44 (2.204-171.43), p=0.008], cardiovascular comorbidities [OR 3.16 (1.32-8.01), p=0.017], hypertension [OR 2.14 (1.04-4.4), p=0.038], active neoplastic disease [OR 3.69 (1.045-1.305), p=0.043] and chronic kidney disease [OR 2.23 (0.897-5.56), p=0.084] were all associated with preexisting diabetes, while comorbid neurodegenerative diseases were associated with COVID-associated hyperglycaemia [OR 3.23 (1.044-10.02), p=0.042]. The median time from symptoms onset to hospital admission was 6 (2-9.5) and 7 (3-10) days for patients with pre-existing diabetes and COVID-associated hyperglycaemia, respectively (p=0.484). Patients with preexisting diabetes and COVID-associated hyperglycaemia reported similar symptoms at the time of hospital admission for COVID-19 pneumonia (Table 1).

**Table 1.** Baseline characteristics of study cohort, according to either preexisting diabetes or COVID-associated hyperglycaemia.

|  | Preexisting diabetes | COVID-associated hyperglycaemia | p | Missing |
| --- | --- | --- | --- | --- |
| N | 121 | 55 |  |  |
| Age, years | 71 (62-77.5) | 69 (54-78) | 0.324 | 0 |
| Sex, male [N (%)] | 84 (71.2) | 37 (63.8) | 0.387 | 0 |
| BMI | 29.16 (26.12-33.8) | 26.52 (23.87-30.12) | 0.014 |  |
| - <25 [N (%)] | 23 (20.5) | 14 (31.8) |  |  |
| - 25-30 [N (%)] | 36 (32.1) | 19 (43.2) | 0.036 | 20 |
| - >30 [N (%)] | 53 (47.3) | 11 (25) |  |  |
| Ethnicity [N (%)] |  |  |  |  |
| - Caucasian | 106 (87.6) | 48 (87.3) |  |  |
| - Hispanic | 7 (5.8) | 3 (5.5) | 0.998 | 0 |
| - Asian | 4 (3.3) | 2 (3.6) |  |  |
| - African | 4 (3.3) | 2 (3.6) |  |  |
| Comorbidities [N (%)] |  |  |  |  |
| - Hypertension | 91 (75.2) | 32 (58.2) | 0.033 |  |
| - Coronary Artery Diseases | 35 (28.9) | 6 (10.9) | 0.012 |  |
| - COPD | 16 (13.2) | 5 (9.1) | 0.617 | 0 |
| - Chronic Kidney Disease | 32 (26.4) | 7 (12.7) | 0.05 |  |
| - Cancer | 23 (19) | 3 (5.5) | 0.021 |  |
| - Neurodegenerative disease | 7 (5.8) | 8 (14.5) | 0.078 |  |
| Preadmission treatment [N (%)] |  |  |  |  |
| - Metformin | 71 (58.7) | 0 |  |  |
| - Sulfonyleureas | 12 (9.9) | 0 |  |  |
| - Repaglinide | 3 (2.5) | 0 |  |  |
| - Acarbose | 3 (2.5) | 0 |  |  |
| - Thiazolidinediones | 1 (0.8) | 0 |  |  |
| - DPP-4 inhibitors | 14 (11.6) | 0 |  |  |
| - GLP-1 agonist | 3 (2.5) | 0 |  |  |
| - SGLT2 inhibitors | 4 (3.3) | 0 |  | 0 |
| - Insulin | 41 (33.9) | 0 |  |  |
| - ASA | 47 (38.8) | 7 (12.7) | <0.001 |  |
| - Statin | 44 (36.4) | 6 (10.9) | 0.001 |  |
| - ACE inhibitors | 28 (23.1) | 11 (20) | 0.699 |  |
| - Angiotensin II R blockers | 28 (23.1) | 6 (10.9) | 0.065 |  |
| - ACEI and/or ARB | 54 (44.6) | 17 (30.9) | 0.099 |  |
| - Calcium channel blockers | 35 (28.9) | 9 (16.4) | 0.091 |  |
| - Beta blockers | 43 (35.5) | 13 (23.6) | 0.162 |  |
| **Admission to the hospital** |  |  |  |  |
| Median time from symptoms to admission, days | 6 (2-9.5) | 7 (3-10) | 0.484 |  |
| Symptoms at onset [N (%)] |  |  |  |  |
| - fever | 93 (78.2) | 46 (85.2) | 0.31 |  |
| - dyspnea | 79 (66.4) | 37 (68.5) | 0.86 |  |
| - cough | 46 (38.7) | 21 (38.9) | 0.99 |  |
| - fatigue/malaise | 35 (29.4) | 10 (18.5) | 0.14 |  |
| - hypo/dysgeusia | 19 (17.3) | 8 (14.8) | 0.82 |  |
| - hypo/anosmia | 17 (14.3) | 8 (14.8) | 0.99 | 0 |
| - myalgia/arthralgia | 19 (16) | 7 (13) | 0.82 |  |
| - headache | 12 (10.1) | 1 (1.9) | 0.066 |  |
| - chest pain | 12 (10.1) | 2 (3.7) | 0.23 |  |
| - diarrhea | 16 (13.4) | 6 (11.1) | 0.81 |  |
| - sore throat | 5 (4.2) | 6 (11.1) | 0.10 |  |
| - vomiting/nausea | 9 (7.6) | 6 (11.1) | 0.56 |  |
| - conjunctivitis | 8 (6.7) | 5 (9.3) | 0.55 |  |
| - abdominal pain | 5 (4.2) | 2 (3.7) | 0.99 |  |
| - skin rash | 2 (1.9) | 0 | 0.99 |  |

### COVID-associated hyperglycaemia was associated with a specific clinical laboratory profile at admission

Upon admission (Table 2), patients with COVID-associated hyperglycaemia exhibited significantly higher white blood cell count [9.7 (6.1-14.3) vs 7.1 (5.4-10.4) ×10^9^/L, p=0.004], neutrophil count [8.4 (4.85-12.9) vs 5.2 (3.9-8.1) ×10^9^/L, p=0.003], and tissue damage markers [LDH 8.04 (5.35-16.63) vs 5.92 (4.42-8.17) μkat/L, p=0.001; AST 0.95 (0.65-1.39) vs 0.63 (0.42-1.14) μkat/L, *p*=0.001; ALT 0.78 (0.47-1.24) vs 0.53 (0.3-0.93) μkat/L, *p*=0.012] compared to patients with preexisting diabetes. Furthermore, an increase in procalcitonin [0.95 (0.49-3.58) vs 0.61 (0.31-1.38) ng/ml, p=0.014] and hemoglobin [12.9 (11.5-14.6) vs 12.5 (10.75-13.5) g/L, p= 0.042] was evident, with a trend also for ferritin [1254 (585-2433) vs 823 (462-1499) μg/L, p= 0.063] and total bilirubin [11.62 (8.16-18.81) vs 9.23 (5.39-14.87) μmol/L, p=0.054]. All the other laboratory variables associated with COVID-19 pneumonia severity, including inflammation indices (CRP and IL-6), hemostatic variables (D-dimer levels, platelet count, fibrinogen, partial thromboplastin time and prothrombin time), biomarkers of myocardial damage (troponin T and Pro-BNP), markers of liver and kidney function (albumin, phosphatase alkaline, creatinine) and glucose control were similar between preexisting diabetes and COVID-associated hyperglycaemia. Antibody responses to SARS-CoV-2 were available for a large subgroup of patients (Table 2). No differences between pre-existing diabetes and COVID-associated hyperglycaemia were evident for IgG, IgM and IgA responses to the SARS-CoV-2 spike protein (RBD or S1+S2), IgG response to NP, autoimmune antibodies anti GAD and anti interferon alpha-4.

**Table 2.**
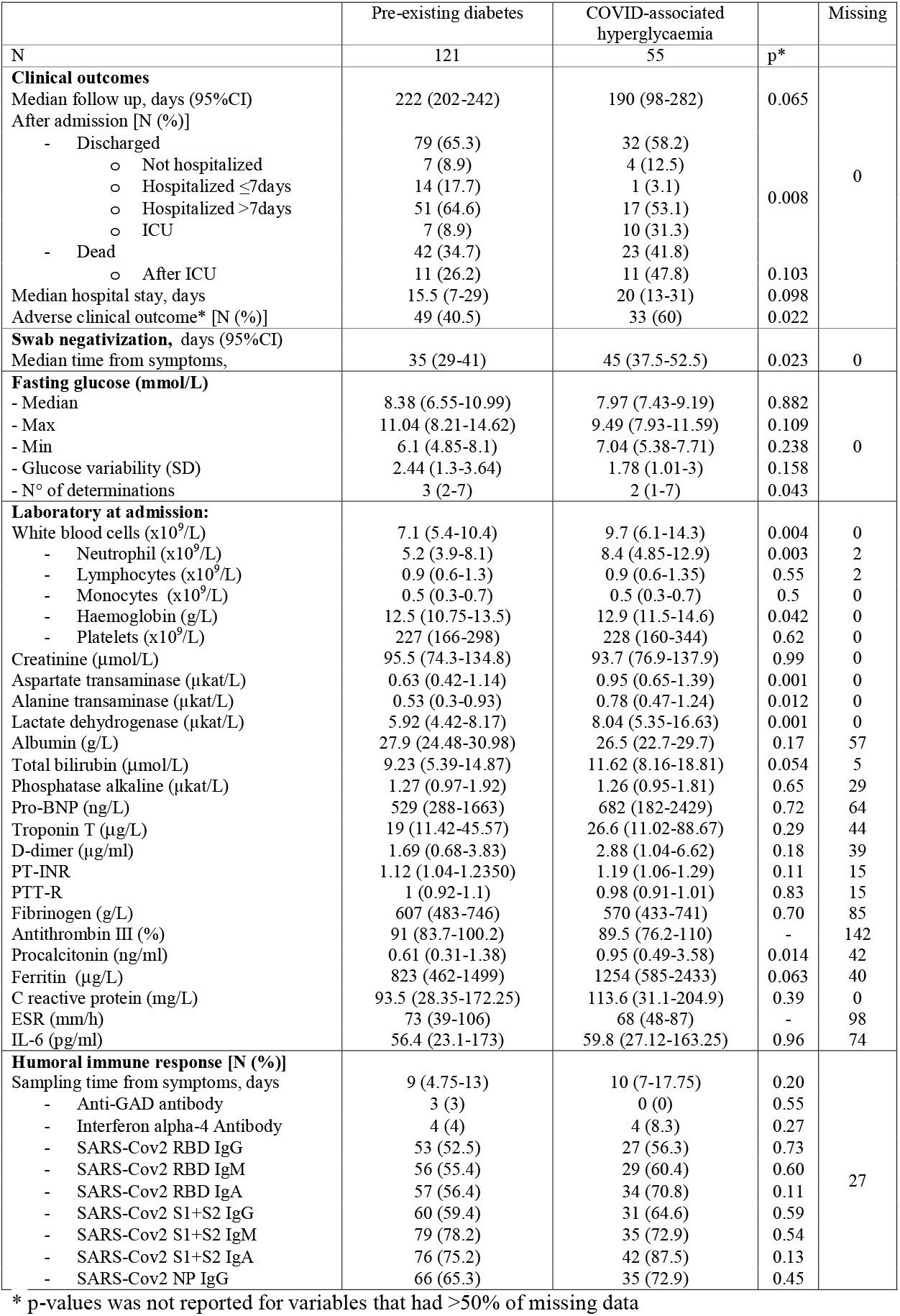
Clinical laboratory profile at hospital admission and clinical outcome.

### COVID-associated hyperglycaemia was associated with a worse clinical outcome

Sex-and age-adjusted Cox proportional hazards model (HR 2.11, CI 1.34-3.31; p=0.001) and Kaplan-Meier estimator log-rank test (p=0.005) indicated that COVID-associated hyperglycaemia was strongly associated with an increased risk of adverse clinical outcome, as defined by composite endpoint of admission to ICU or death, whichever occurred first. (Fig 1 and Table 2). Laboratory variables associated with adverse clinical outcome in univariate Cox proportional hazards model, included inflammation indices (CRP, IL-6, ferritin, procalcitonin, low lymphocytes count, high white blood cell and neutrophil count), hemostatic parameters (D-dimer levels, fibrinogen and partial thromboplastin time), biomarkers of myocardial damage (troponin T and Pro-BNP), markers of liver and kidney dysfunction (albumin, total bilirubin and creatinine), markers of tissue damage (lactate dehydrogenase and aspartate transaminase) and markers of glucose control (fasting plasma glucose and glucose variability). Of note, the presence of hypertension (HR 0.59, CI 0.36-0.94; p=0.028) and the treatment with Angiotensin II receptor blocker (HR 0.44, CI 0.22-0.85; p=0.014) was associated to a decreased risk for adverse clinical outcome. A multivariable analysis using two different models confirmed COVID-associated hyperglycaemia (HR 2.68, CI 1.47-4.89; p=0.001 and HR 1.72, CI 0.95-3.09; p=0.073 respectively) as an independent predictor of adverse clinical outcome (Figure 1B) for patients with COVID-19 pneumonia. To evaluate the specific impact of clinical characteristics and laboratory tests, the univariate Cox proportional hazards model was also conducted separately for patients with preexisting diabetes and those with COVID-associated hyperglycaemia. Although many analysed factors were similarly associated with COVID-19 adverse clinical outcome in either COVID-associated hyperglycaemia or preexisting diabetes, some qualitative or quantitative differences emerged (fig 1B).

**Figure 1.**
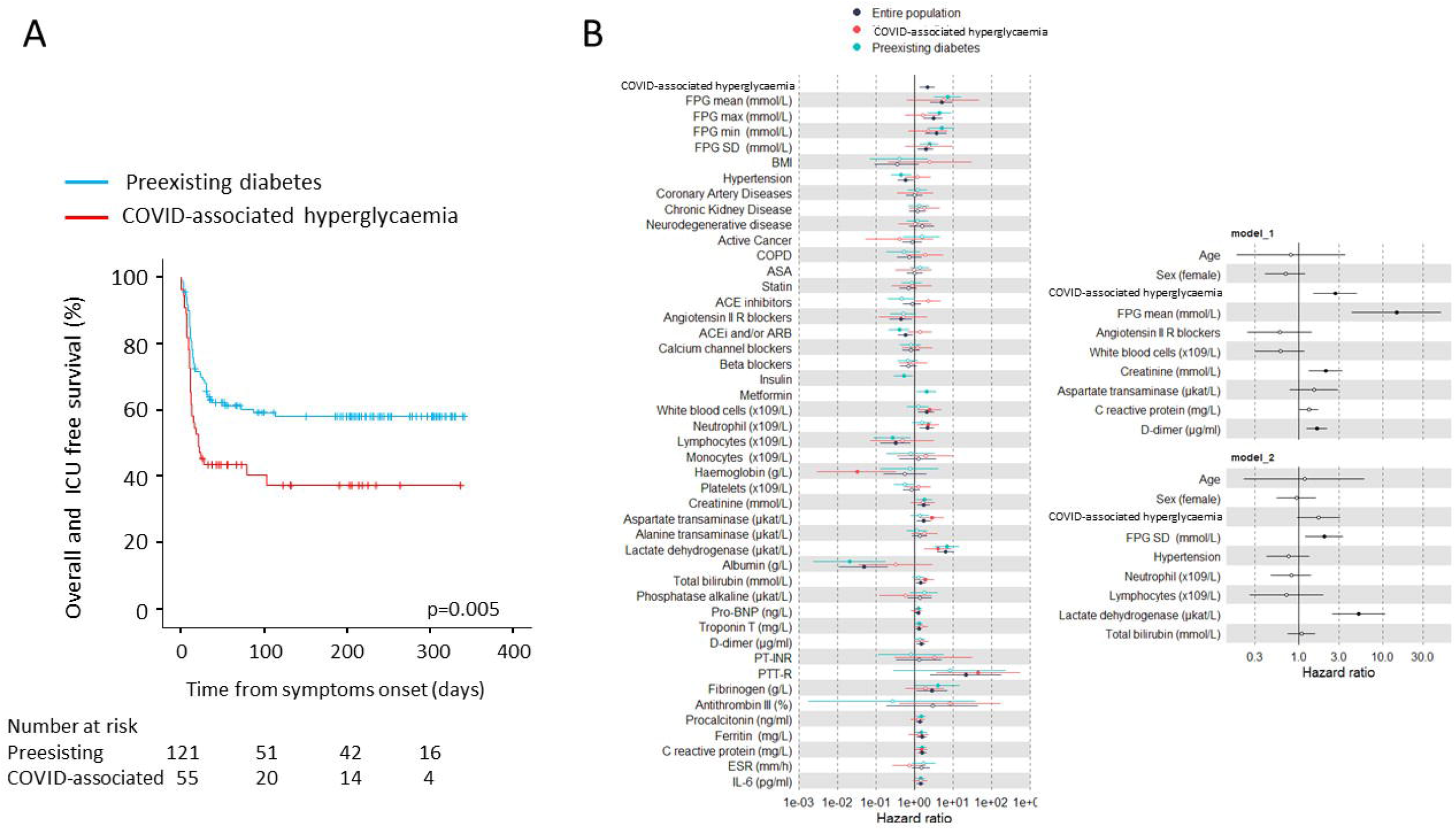
a-b. Adverse clinical outcome in patients with Sars-CoV-2 infection with either preexisting diabetes or COVID-associated hyperglycaemia. Kaplan-Meier estimates of survival without adverse clinical outcome for 176 patients with COVID-19 pneumonia (panel a). Survival without adverse clinical outcome (defined by composite endpoint of admission to ICU or death, whichever occurred first) was estimated for patients with COVID-associated hyperglycaemia (n=55) or preexisting diabetes (n=121). The log-rank test was used to test differences between the two groups. Crosses indicate censored patients (censoring for death or end of follow-up). The forest plots (panel b) show the hazard ratios for survival without adverse clinical outcome according to presence of new-onset or preexisting diabetes. Left panel: univariate Cox regression analysis adjusted for sex and age. Right panel: multivariate Cox regression analysis adjusted for sex and age, including variables significant at the level of <0.1 in the univariate analysis. Variables were excluded from multivariate Cox regression if they showed substantial biological redundancy with other variables (e.g. aspartate aminotransferase vs alanine aminotransferase) or had data obtained for fewer than 78% of patients. Dots represent the HR, lines represent 95% confidence interval (CI), and solid dots indicate P < 0.05.

**Figure 2.**
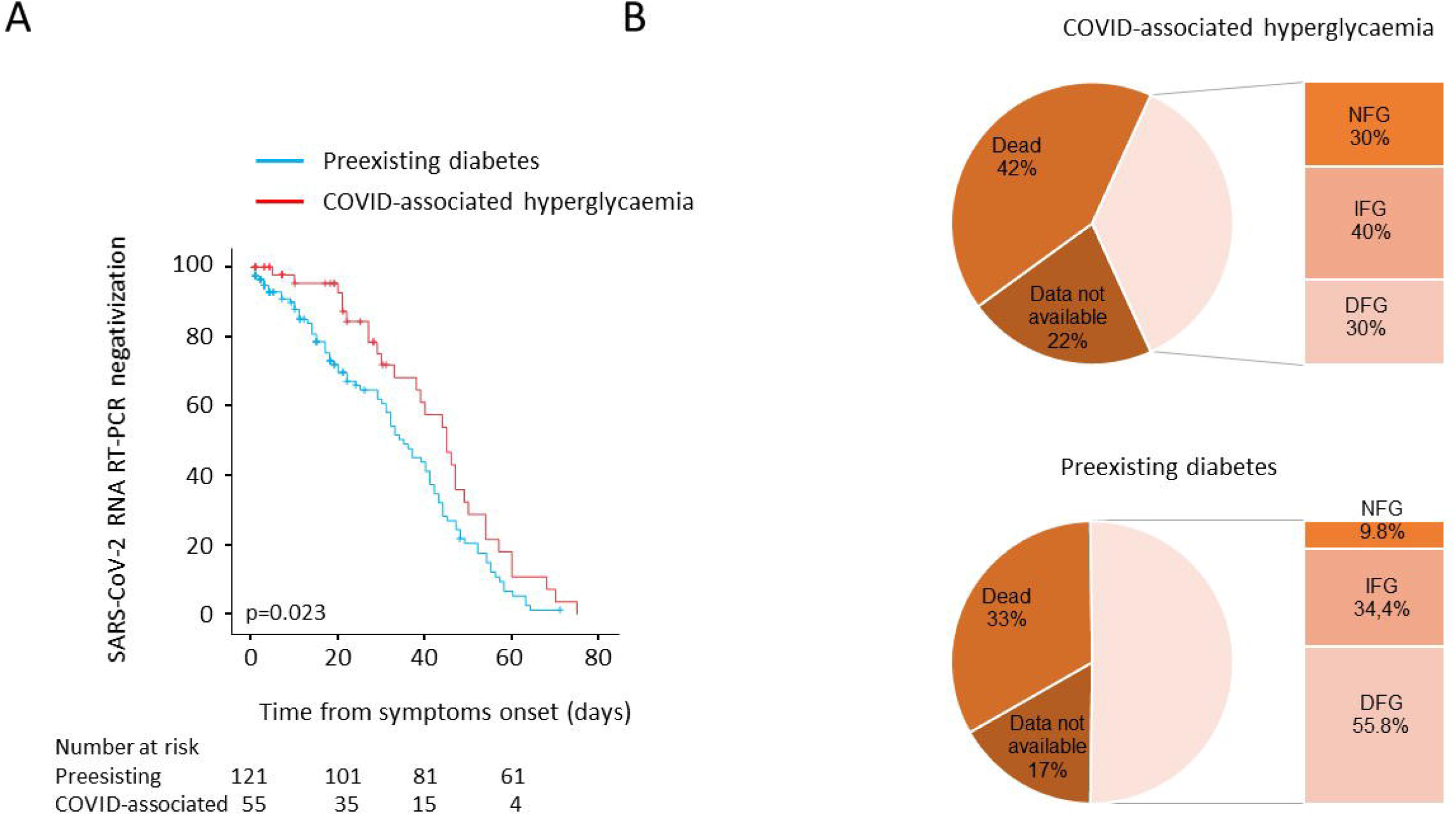
a-b. Post discharge follow-up: time to NT swab negativization and persistence of hyperglycemia. Kaplan-Meier estimates of the time to NT swab negativization for 176 patients with COVID-19 pneumonia (panel a). NT Swab negativization was estimated in patients with COVID-associated hyperglycaemia (n=55) or preexisting diabetes (n=121). The log-rank test was used to test differences between groups. Crosses indicate censored patients (censoring for death or end of follow-up). Fasting blood glucose during the post discharge follow up (panel b). Valid glucose measurements during follow up (median 6 month) was available for 61 out of 121 and 20 out of 55 patients with preexisting diabetes or COVID-associated hyperglycaemia, respectively. NFG: Normal Fasting Glucose, <5.6 mmol/L; IFG: Impaired Fasting Glucose 5.6-6.9 mmol/L; DFG: Diabetes Fasting Glucose ≥ **7** mmol/L.

### Post discharge follow-up: time to NT swab negativization and persistence of hyperglycemia

As the prolonged viral shedding is a risk factor for poor outcome of COVID-19 and may interfere with the immunological mechanism of virus elimination, in our study population we evaluated the time to NT swab negativization. Sex-and age-adjusted Cox proportional hazards model (HR 0.661, 95% CI 0.43-1.02; p=0.063) and Kaplan-Meier estimator log-rank test (45 days, 95% CI 37-52 vs 35 days, 95% CI 29-41; p=0.023) documented a negative association between COVID-associated hyperglycaemia and the time to NT swab negativization. We also evaluated fasting blood glucose during the post-discharge follow up (i.e., at the 1, 3, 6, and 9 month outpatient visits). Among 121 subjects with preexisting diabetes, 40 died and 20 had no glucose measurements during follow up (median 6 month). Of the remaining 61 patients with a prescribed treatment for diabetes, six had normal fasting glucose (NFG) [9.8%, 90 (84.5-91.75) mg/dl], 21 impaired fasting glucose (IFG) [34.4%, 112 (108-119) mg/dl] and 34 fasting glucose in the diabetes range (DFG) [55.7%, 160.5 (133-189.5) mg/dl]. Among 55 patients with COVID-associated hyperglycaemia, 23 died and 12 had no glucose measurements during follow up (median 6 month). Of the remaining 20 patients, none had a prescribed diabetes treatment during follow up: six showed normalization of their fasting glucose [30%; 86 (84-90 mg/dl)], while 14 were confirmed as having either IFG [n=8, 40%, 104 (100.2-111)] or DFG [n=6, 30%; 136 (126-148) mg/dl].

## Discussion

Hyperglycaemia [5] is emerging as a common feature among patients hospitalized for COVID-19 pneumonia [22]. This COVID-associated hyperglycaemia still needs to be adequately characterized since there are only few studies documenting how it differs from preexisting diabetes in COVID-19 patients. To address this issue, we studied a cohort of 176 adult patients with COVID-19 pneumonia with a diagnosis of preexisting diabetes or hyperglycaemia. Our study generated several interesting findings. First, patients with COVID-associated hyperglycaemia had significantly less comorbidities related to the metabolic syndrome than those with preexisting diabetes. The main reason for this difference probably lies in the lower prevalence of obesity and a shorter duration of hyperglycemia. Consistent with this, prior to admission only a low proportion of patients was prescribed antihypertensive drugs, lipid-lowering agents or antiplatelet therapy. However, it cannot be excluded that the lower prevalence is an effect related to an underdiagnosis driven by the absence of diabetes diagnosis. Similarly, the higher prevalence of neurodegenerative disease in subjects with COVID-associated hyperglycaemia could be related with lower utilization of health care and therefore lower rates of diagnosis of diabetes pre admission in patients with a cognitive impairment. Second, patients with COVID-associated hyperglycaemia showed increased levels of inflammatory markers and indicators of multi-organ injury compared to patients with preexisting diabetes. Third, the prevalence of autoimmune biomarkers as anti GAD (a marker of islet autoimmunity) or interferon alpha-4 antibody (an autoimmune marker recently associated with COVID-19 severity) [23] was generally very low and did not differ between COVID-associated hyperglycaemia and preexisting diabetes. Fourth, COVID-associated hyperglycaemia was associated with a poorer clinical outcome and a delayed time to viral clearance compared to preexisting diabetes, despite the presence of a superimposable humoral responses to SARS-CoV-2 and similar glucose levels. Fifth, COVID-associated hyperglycaemia reversed in most patients when the viral infection resolved.

Whether COVID-asociated hyperglycaemia should be considered a specific clinical entity is a matter of discussion. In terms of pathophysiology, hyperglycaemia may include (a) “stress-induced” hyperglycemia; (b) previously unrecognized (pre)diabetes (either type 2 or, less likely, type 1 diabetes); (c) a form of diabetes due to the direct effect of the SARS-CoV-2 virus on the pancreas; or (d) a form of diabetes secondary to the treatment of COVID-19 (e.g., with glucocorticoids or other medications possibly inducing secondary diabetes) [24-31]. To adequately addressing this question would require the availability of pancreatic tissue or advanced studies on insulin secretion and resistance, which would be unrealistic to perform during the acute phase of COVID-19 pneumonia. Moreover, it is very likely that more than one cause may contribute to the hyperglycemia associated with COVID. In our study, the absence of markers of islet autoimmunity and the lower prevalence of comorbidities related to metabolic syndrome do suggest specific pathophysiological mechanisms responsible for hyperglycaemia. More specifically, the extremely low prevalence of GAD autoantibodies excludes a previously unrecognized autoimmune (pre)diabetes (i.e. LADA) as possible cause of new onset diabetes. The use of glucocorticoids in the first wave of the pandemic was very limited and, therefore, unlikely to have played a role in the pathogenesis of hyperglycaemia. Moreover, as hyperglycaemia reverted in most patients when the infection resolved, it is reasonable to hypothesize that reversible transient factors, such as inflammation-induced insulin resistance, may play a role in causing hyperglycemia in those patients [28].

Regardless of the pathophysiological mechanism, our study documented that COVID-associated hyperglycaemia was associated with an adverse clinical outcome of SARS-CoV-2 infection and that this association was independent from the major risk factors for disease severity. As good glycemic control was indeed shown to reduce disease severity and mortality in COVID-19 patients with hyperglycemia[32], early recognition and treatment of COVID-associated hyperglycaemia may greatly benefit these patients. Therefore, our findings do strongly support the need to screen all patients with COVID-19 pneumonia for hyperglycemia (i.e., blood glucose and/or HbA1c) at the time of admission, despite a mute personal or family history of diabetes.

Since blood glucose levels in our study were not different in patients with either COVID-associated hyperglycaemia or preexisting diabetes, factors other than hyperglycemia should be considered to explain the differences in clinical outcome. A role could be played by a different drug treatment prior to admission. Of note, in our study sex-and age-adjusted Cox proportional hazards model indicated that angiotensin-converting enzyme inhibitors (ACEi) and/or angiotensin receptor blockers (ARB) treatment was significantly associated with a better clinical outcome in preexisting diabetes. Most studies, including several meta-analyses, found no substantial differences in the risk for severe COVID-19 pneumonia associated with the prescription of common classes of antihypertensive medications[33]. On the other hand, evidence of the beneficial effects of chronic ACEi/ARB use, especially in hypertensive cohort of patients with COVID-19 pneumonia, was reported[34-37]. As the prevalence of hypertension is higher among patients with preexisting diabetes, ACEi/ARB use may mediate a selective beneficial effect. Similarly, in our study around 60% of patients with preexisting diabetes was taking metformin and the treatment was associated with a better clinical outcome. Concordantly, previous observational studies suggested that in patients taking metformin the risk for COVID-19 mortality is reduced[38, 39]. Of great interest is also the finding of a slow viral clearance in patients with COVID-associated hyperglycaemia. Discordant data exist about differences in viral clearance or shedding in people with diabetes [40]. As increased glucose levels and glycolysis promoted Sars-CoV-2 replication in monocytes via ROS/HIFα pathway activation leading to secondary T-cell dysfunction [41], this could explain the delayed viral clearance associated with the acute hypeglycaemia in patients with COVID-associated hyperglycaemia.

Our study has some limitations. First, the analysis was performed on a subcohort of 176 patients selected for having hyperglycemia or preexisting diabetes out of 584 subjects of our original cohort (12). All patients with diabetes/hyperglycaemia were included, and as for age, sex, hyperglycaemia prevalence, and the clinical outcome of our cohort appears superimposable to those reported by many authors. Despite this, we cannot exclude a selection bias. Second, our cohort is limited to hospitalized patients with COVID-19 pneumonia. Third, we only assessed fasting blood glucose and we acknowledge that more specific indicators of beta cell function, such as serum insulin and or C peptide levels, should have been measured. Fourth, the lack of HbA1c measurements in our cohort is a limitation. The evidence of normal HbA1c at the time of admission would indicate no history of hyperglycemia and confirm the diagnosis of COVID-associated hyperglycaemia. HbA1c levels were available for some patients with preexisting diabetes (51 (41-58) mmol/mol; 6.8%), but in very few patients with COVID-associated hyperglycaemia (38 (36-40) mmol/mol; 5.6%), and for this reason were not included in the analysis.

In conclusion, COVID-associated hyperglycaemia is emerging as a complication of Sars-CoV-2 infection and this clinical entity still needs to be adequately characterized in comparison with preexisting diabetes. It is clear from our study that patients with COVID-associated hyperglycaemia had increased levels of inflammatory markers and indicators of organ injuries associated with a poorer clinical outcome compared to preexisting diabetes. This strongly support the need to screen all COVID-19 patients for hyperglycemia at the time of admission, despite a mute personal or family history of diabetes, and to treat them in order to reach and maintain a good glycemic control during the hospitalization for COVID-19 pneumonia.

## Data Availability

Some or all datasets generated during and/or analyzed during the current study are not publicly available but are available from the corresponding author on reasonable request.

## DECLARATIONS

Professor Lorenzo Piemonti is the guarantor of this work and, as such, had full access to all the data presented in the study and takes responsibility for the integrity of data and the accuracy of data analysis.

### Availability of data

The datasets generated during and/or analyzed during the current study are not publicly available due to concerns about data confidentiality, but are available from the corresponding author on reasonable request.

### Individual contributions

LP, VL and MS contributed to the conception of the study, wrote the manuscript, researched data and contributed to the discussion. AL, AC and CM, contributed to the acquisition, analysis and interpretation of clinical data, contributed to the design of the study and critically reviewed/edited the manuscript. PRQ and GL contributed to the collection of biological samples, managed the biobanking activities, critically revised the manuscript, recruited patients and managed sample biobanking.

### Ethics approval

All procedures performed in studies involving human participants were in accordance with the ethical standards of the institutional and/or national research committee and with the 1964 Helsinki Declaration and its later amendments or comparable ethical standards. The study was approved by the Ethics Committee of the IRCCS Ospedale San Raffaele (protocol number 34/int/2020).

### Consent to participate

Informed consent was obtained from all individual participants included in the study.

## Acknowledgments

This work was funded by IRCCS Ospedale San Raffaele (Program Project COVID-19) and Ministero della Salute (COVID-2020-12371617).

